# Penetrance of pathogenic epilepsy variants is low and shaped by common genetic background

**DOI:** 10.1101/2025.10.16.25338116

**Authors:** Remi Stevelink, Martijn Piet, Yorgos Bos, Ruben van ’t Slot, Kees P.J. Braun, Bobby P.C. Koeleman

## Abstract

The identification of pathogenic variants in developmental epileptic encephalopathy (DEE) genes can be vital for counselling and individualized treatment. Penetrance is usually considered to be high or full, although this has never been studied in population cohorts. Recent evidence shows that common polygenic risk factors (PRS) are increased in DEE cases, suggesting it might modify risk. Here, we calculated the penetrance of autosomal dominant epilepsy variants that have previously been classified as (likely) pathogenic in ClinVar, in two large cohorts (n=42,863 and n=386,306) and assessed whether common variant PRS could modify risk. Most people carrying pathogenic DEE variants did not have epilepsy. Penetrance estimates suggested that the probability of epilepsy in pathogenic variant carriers ranges between 4.1% and 9.8%. Among people carrying epilepsy variants, PRS was predictive of an epilepsy diagnosis. A high PRS was associated with increased risk of severe epilepsy whereas a low PRS seems protective in people carrying a pathogenic variant. Our results suggest that average variant penetrance is lower than expected and modified by PRS. A high PRS combined with a pathogenic variant may be necessary to develop a severe epilepsy phenotype like DEE. Reconsidering penetrance assumptions could improve variant classification and diagnostic yield. Our findings may enhance genetic counselling by refining risk estimates and could extend to other diseases.

## Introduction

Since 1995, over a thousand epilepsy genes have been discovered.^1^ Diagnosing a pathogenic variant as etiological cause of epilepsy is important to determine prognosis and enable targeted treatment.^2^ Finding a pathogenic variant in a child with developmental epileptic encephalopathies (DEE) is generally deemed sufficient to diagnose ‘monogenic epilepsy’.^3,4^

Surprisingly, recent studies show that individuals with DEE and a causal pathogenic variant also carry an increased burden of common epilepsy risk variants, quantified as polygenic risk scores (PRS).^5–8^ However, it remains unclear whether people who carry a pathogenic variant and a low PRS remain healthy or show a milder phenotype.

Although reports of incomplete penetrance are documented (e.g. in *SCN1A, SCN1B* and *GABRG2*),^6,9^ autosomal dominant DEE variants are generally thought to be of high or full penetrance, as they are rarely inherited from a healthy parent.^3,4^ Many DEE genes have been discovered by selecting for *de novo* variants that occur in multiple severely affected patients, but it is unknown whether these variants also occur in people with mild epilepsies or healthy individuals. Penetrance of recurrent variants is usually determined within large epilepsy families,^6,10^ however, these estimates could be inflated since family members share environmental factors and polygenic background that are unrelated to the pathogenic variant itself.

Variants are often considered benign if they are reported in multiple healthy subjects, since individual pathogenic epilepsy variants are exceedingly rare in population databases such as GnomAD or ExAc.^11,12^ However, there are thousands of established pathogenic epilepsy variants and their collective population prevalence and penetrance remains elusive.

In this study, we first estimated the penetrance of autosomal dominant DEE variants by assessing their prevalence in large cohorts of unrelated people with and without epilepsy. Next, we calculated their PRS to assess whether common risk variants could modify penetrance.

## Materials & methods

### Study population

We used genomic and phenotypic data from two large cohorts: Genomics England (GEL) and the UK Biobank (UKB).

The GEL served as a discovery cohort. Around 78.000 patients with rare diseases, cancer and unaffected relatives were recruited via centres across the United Kingdom at a mean age of 30 years (range 0-99).^13,14^ Detailed standardised phenotypic information including disease status was collected for all subjects. Informed consent was obtained from all subjects, parents or legal guardians.

The UKB was used for replication. This population cohort included 500,000 individuals aged 40–69 at recruitment.^15^ Subjects were recruited irrespective of diseases, socioeconomic status or ethnicity. Mental capacity to provide informed consent was assessed, likely resulting in exclusion of profound intellectual disability.

Subjects of both cohorts underwent germline whole genome sequencing (WGS) using Illumina next generation sequencing with a mean depth of 32x.

### Phenotype definition

In GEL, epilepsy cases were identified if they were recruited under the category ‘Early onset or syndromic epilepsy’, which primarily constituted DEE and epilepsy plus other features like intellectual disability, diagnosed by a neurologist. To find additional subjects not specifically recruited for epilepsy, we also used ICD codes to define cases with epilepsy (ICD G40), which we refined by excluding acquired structural causes (ICD codes C90-96, C71, S06, I60-63). Similar to a previous study, we defined ‘severe epilepsy’ when people were recruited for epileptic encephalopathy, epilepsy plus intellectual disability, or had epilepsy plus an ICD diagnosis of intellectual disability (F70-F79) or disorder of psychological development (F80-89).^5^ Controls had no epilepsy diagnosis (by recruitment or ICD code). No subjects were specifically recruited for epilepsy in the UKB, therefore, only the above ICD definitions were used to classify cases and controls, restricted to age at epilepsy diagnosis prior to 18 years of age, under the assumption that monogenic epilepsy would manifest in childhood.^16^ Year of diagnosis was inconsistently reported in GEL and therefore not used.

### Selection of autosomal dominant pathogenic DEE variants

Similar to penetrance studies in other diseases,^17,18^ we queried the ClinVar database (version 20240708) for coding or splicing variants in autosomal dominant DEE genes (Genes4Epilepsy v2024-3),^1^ retaining only variants classified as pathogenic, likely pathogenic, or pathogenic/likely pathogenic (hereafter referred to as ‘pathogenic epilepsy variants’ for brevity). We removed variants with conflicting evidence or annotations suggesting incomplete penetrance (e.g. “low penetration”, “likely risk allele”, “risk factor”). Since variants in epilepsy genes can be associated with different diseases, we further filtered variants for a disease association with epilepsy, yielding in a list of 4294 pathogenic variants across 92 DEE genes.Variants were filtered on a minor allele frequency <0.0001 in each cohort. Finally, we performed manual curation of each identified variant to assess evidence of pathogenicity and removed variants where the predicted variant effect is not a known disease mechanism and/or where the specific variant is not robustly associated with epilepsy.

### Subject quality control

To avoid inflation of PRS and penetrance estimates due to shared polygenic and environmental factors, we filtered both cohorts to include only unrelated subjects using genetically inferred relatedness (KING relatedness pruning using a threshold of >0.0422, corresponding to 3^rd^ degree relatedness or higher). European-ancestry individuals were selected using genetic principal component analyses to increase homogeneity and enable PRS analyses using summary statistics from the largest European-only epilepsy GWAS.^19^ Standard quality control measures were applied in both GEL and UKB, including exclusion of duplicates, outliers based on missingness, and genotyping heterozygosity, as described previously.^13,15^

### Polygenic risk score analysis

PRS were calculated with PRSice-2^20^ using summary statistics from the largest European all epilepsy GWAS containing 27,559 epilepsy cases.^19^ WGS data was filtered on minor allele frequency >0.01, Hardy-Weinberg disequilibrium (p<1e-10 in GEL and p<1e-54 in UKB) and >98% call rate per single nucleotide polymorphism (SNP). SNPs were pruned to a subset of uncorrelated SNPs (R^2^<0.1) and SNPs with P<0.5 were selected based on optimisation in a previous study.^21^ Individual PRS were than calculated as the sum of weighted effect alleles, after which PRS were standardized using Z-score transformation: 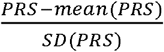.

### Statistical analysis

Logistic regressions were used to perform PRS group comparisons, corrected for the first four principal components of ancestry and sex, as described earlier.^21^

We used linear regression analyses corrected for age and sex to compare fluid intelligence scores between groups in the UKB cohort.

Chi-squared tests were used to compare pathogenic variant prevalence between the GEL and UKB cohorts.

To assess penetrance within the GEL cohort, we assessed the proportion of people with epilepsy amongst subjects who carried a pathogenic DEE variant. We corrected for ascertainment bias by correction penetrance estimates using the following formula as described previously:^22^

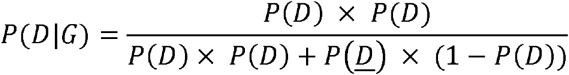

where D=disease, G=genotype (carrying a pathogenic variant), and 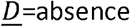 of disease. P(D|G) is the penetrance (probability of disease given a genotype); P(G|D) is the genotype frequency in cases; 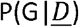 is the genotype frequency in controls and P(D) the general population prevalence of epilepsy. We assumed a 0.7% prevalence of broadly defined epilepsy and 0.29% prevalence of severe epilepsy phenotypes (DEE and epilepsy plus developmental impairment), based on recent studies.^16,23^

All statistical tests were performed using R statistical software (version 4.3.2) and two-tailed P-values are reported.

## Data availability

This project was conducted using publicly available data and software. Genetic and phenotypic data from the UK Biobank can be applied for at https://www.ukbiobank.ac.uk/enable-your-research/register. Access to data from GEL can be applied for at https://research.genomicsengland.co.uk/application/. PRSice-2 is available at https://choishingwan.github.io/PRSice/.

## Results

### Penetrance of pathogenic epilepsy variants in GEL

Among 42,863 individuals in the GEL discovery cohort, we identified 45 carriers (0.10%) of a (likely) pathogenic autosomal dominant epilepsy variant. Of the 45 carriers, 14 (31%) had epilepsy, 13 of whom had a severe epilepsy phenotype; one subject had genetic generalised epilepsy with normal development. Among the 31 variant carriers without epilepsy, two (6%) had developmental impairment. The average age of people without epilepsy at last follow-up was 50 years (range 15-87), by which a monogenic epilepsy should have already manifested.^16^

Variants were distributed across 15 genes; 5 variants were found in multiple people, while 32 (86%) were singletons (see **Supplementary Table 1** for a list of all variants and annotations). Regarding variant types, 23 (51%) of subjects carried a missense variant, 11 (24%) a nonsense variant, 4 (9%) a frameshift variant and 7 (16%) an intronic/splice variant. There was no difference in the proportion of missense variants between epilepsy cases and controls who carried pathogenic variants (OR=0.62; p=0.59).

Using a broad definition of epilepsy, 1,839 individuals in GEL (4.3%) had a diagnosis of epilepsy. This yields an ascertainment-adjusted penetrance estimate of 6.8%, assuming a general population prevalence of 0.7%.^23^ Penetrance of nonsense and frameshifts variants was 7.5%. Applying a stricter definition of severe epilepsy (DEE or epilepsy with developmental impairment), we found 1117 cases, corresponding to a penetrance of 4.1%, assuming a 0.29% population prevalence of severe epilepsies.^16^ Thus, carrying a pathogenic variant was associated with a penetrance of 6.8% for any epilepsy and 4.1% for severe epilepsy, representing relative risks of 9.7 and 14.1, respectively.

### Replication in UKB

In the UKB, 297 of 392,123 individuals (0.076%) carried a pathogenic epilepsy variant. This frequency was lower than in GEL (Chi-square=4.21, p=0.040), but similar to GEL controls after excluding epilepsy cases (Chi-square=0.0001, p=0.99). Nine of the people carrying a pathogenic variant had an ICD diagnosis of epilepsy (3.0%), none of which had a severe epilepsy phenotype (DEE or epilepsy plus developmental impairment). None of the unaffected carriers had intellectual disability. Fluid intelligence scores were compared between unaffected carriers and controls to assess potential subclinical or undiagnosed intellectual disability: scores were not significantly different between the groups (beta= - 0.32, p=0.054; **Supplementary Fig. 1**).

Pathogenic variants in UKB were distributed across 34 genes, 13 of which overlapped with those found in GEL. Thirty-four variants were found in multiple individuals, while 78 (70%) were singletons (**Supplementary Table 2**) When assessing variant types, 211 subjects carried a missense (71%), 44 a nonsense (15%), 11 a frameshift (4%), and 315 an intronic/splice (10%) variant. As in GEL, there was no difference in the proportion of missense variants between epilepsy cases and controls who carried pathogenic variants (OR=0.80; p=0.77).

Using a broad epilepsy definition, 796 individuals in UKB (0.2%) had a diagnosis of epilepsy, yielding a penetrance estimate of 9.8% and a relative risk of 14 (vs. the general population prevalence of 0.7%). Among carriers of nonsense or frameshift variants, penetrance was 16.8% (3 cases, 52 controls). Penetrance estimates represent the average across all variants; most variants were singletons and only 4 identical variants were found in cases and controls across GEL and UKB.

### PRS as modifier of penetrance and severity

We assessed whether common epilepsy risk variants, quantified as PRS, modify epilepsy variant penetrance. Among people carrying a pathogenic variant, individuals with epilepsy had significantly higher PRS compared to people without epilepsy (OR=1.84, p=0.047; **Fig. 1A**). Subjects in the highest third of the PRS distribution had a 8-fold higher risk of having epilepsy compared to the lowest quantile (OR=8.1; p=0.04; **Fig. 2**). The area under the receiver operating characteristic curve (AUC) for distinguishing epilepsy cases from non-cases among pathogenic variant carriers was 0.80 (95%CI 0.67-0.93; **Fig. 1B**), suggesting fair predictive ability. We then compared PRS values across groups. Among individuals with epilepsy, those with a pathogenic variant had similar PRS to those without a pathogenic variant (OR=0.80; p=0.38). Unaffected variant carriers had a non-significantly lower PRS compared to controls without a pathogenic variant (OR=0.72; p=0.069). Epilepsy cases with and without a pathogenic variant had a higher PRS compared to controls without a variant (OR=1.91; p=0.019 and OR=1.43; p<2×10^-16^, respectively; **Fig. 3A**; see **Supplementary Table 3** for statistics of all group comparisons).

**Figure 1:**
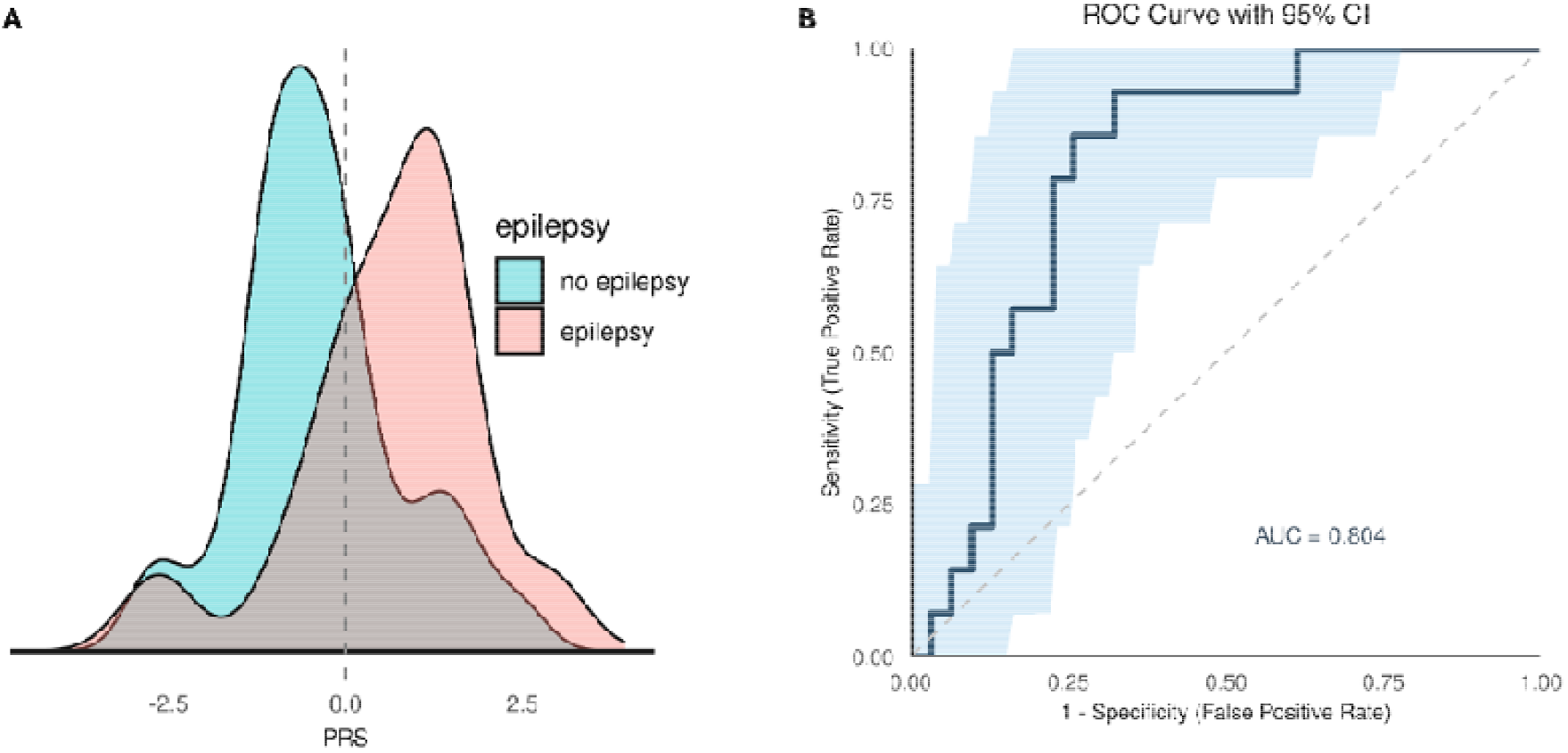
PRS to predict epilepsy among people carrying pathogenic variants. (A) Normal density distribution of epilepsy PRS is plotted in cases (red) and controls (blue) among carriers of pathogenic epilepsy variants. (B) Sensitivity and 1-specificitity with 95% confidence interval shaded in blue are plotted, with the area under the receiver operator curve (AUC) displayed below.

**Figure 2:**
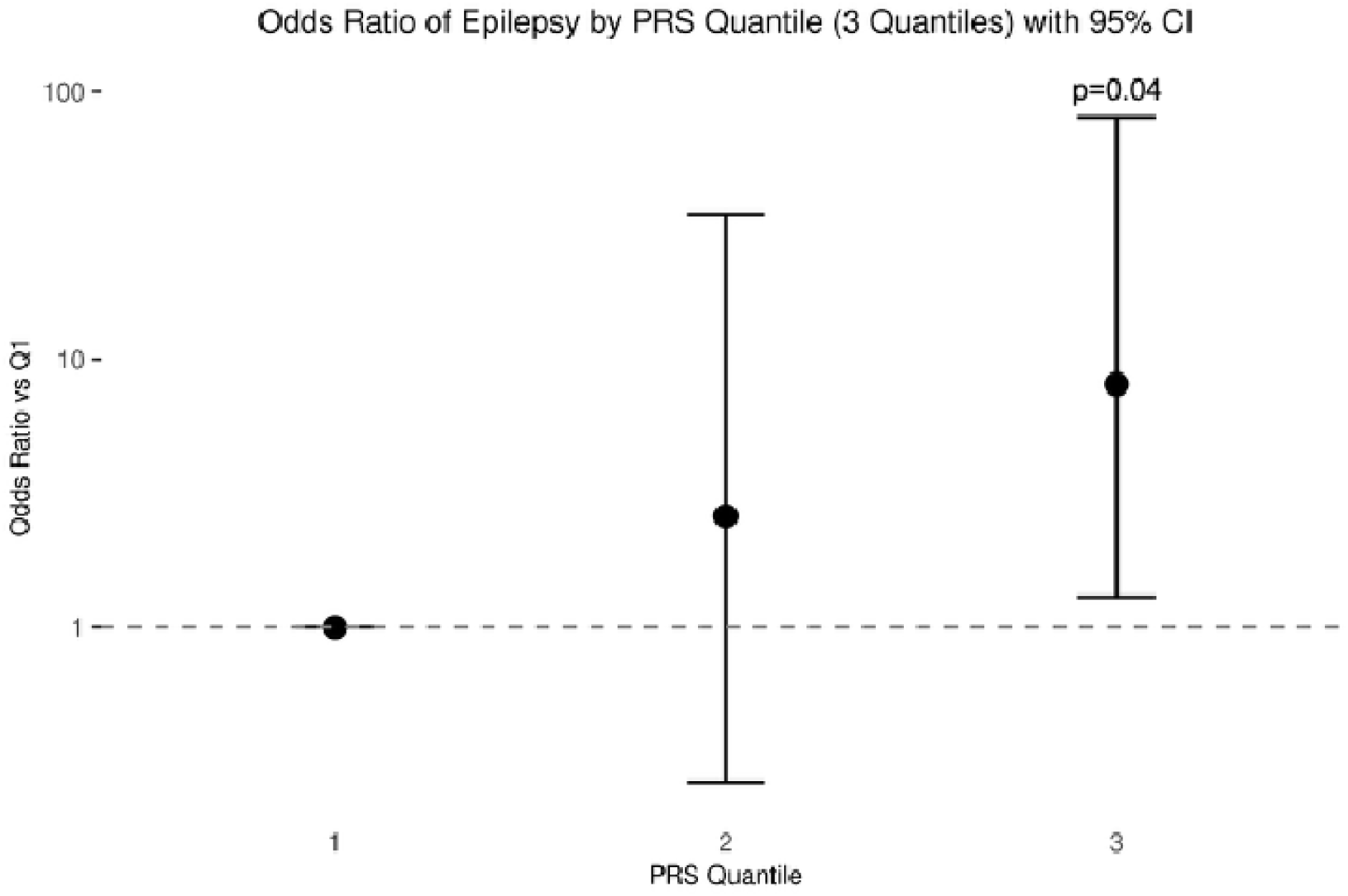
Comparison of epilepsy risk among people carrying pathogenic variants stratified by PRS quantile. Subjects in the GEL cohort are split in three equal quantiles, after which logistic regression is performed to assess epilepsy risk, quantified as odds ratio (visualised on a 10-logarithmic scale for clarity), compared to the first quantile. Whiskers represent 95% confidence intervals.

**Figure 3:**
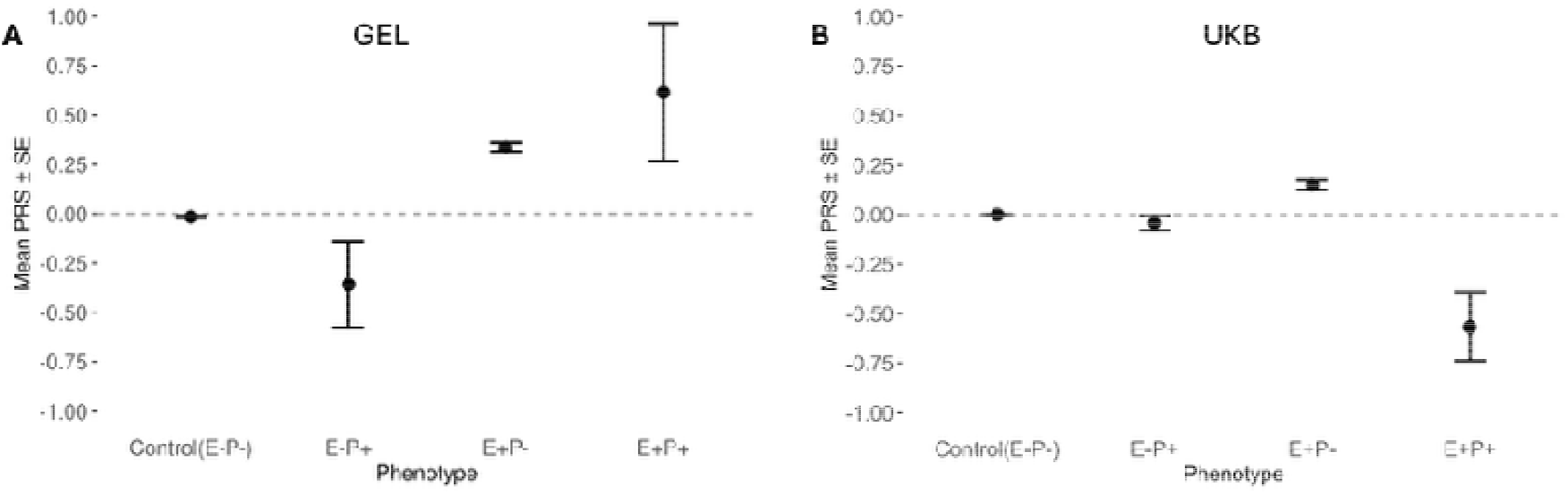
Epilepsy PRS compared between people with and without epilepsy (E), stratified by pathogenic variant status (P). (A) Mean PRS scores in the GEL cohort (including primarily severe epilepsies) are displayed with standard errors as whiskers. (B) PRS scores in the UKB replication cohort (depleted from severe epilepsies)

Replication in the UKB cohort showed no significant PRS difference between epilepsy cases and unaffected controls who both carried a pathogenic variant (OR=0.35; p=0.086; **Fig. 3B**). Within the UKB, epilepsy cases with a pathogenic variant had a lower PRS than both controls (OR=0.33; p=0.032) and epilepsy cases without a pathogenic variant (OR=0.20; p=0.007). Unaffected variant carriers also had a non-significantly lower PRS compared to controls without a pathogenic variant (OR=0.87; p=0.12; see **Supplementary Table 4** for statistics of all group comparisons).

## Discussion

Our results suggest that the penetrance of pathogenic epilepsy variants may be lower than commonly assumed and modified by common risk variants. This has important implications for the interpretation of variant pathogenicity. Under the assumption of high or full penetrance, it is common practice to label variants as benign or of uncertain significance if they are present in multiple unaffected population controls, or in unaffected parents when using a trio-WES approach.^11,12^ However, our study’s low average penetrance warrants caution to label variants as benign when unaffected family members or population controls harbour the same variant. Conversely, finding a pathogenic variant might be insufficient to diagnose a specific epilepsy syndrome, especially without considering the combined effect of common epilepsy risk variants. Potentially, a wide range of variants might be reclassified if assumptions about penetrance are reconsidered to improve diagnostic yield. Furthermore, our findings could have relevance for genetic counselling in carriers of established pathogenic variants, where incorporating polygenic risk might refine risk estimates for their children.

Our results suggest that PRS modifies penetrance of pathogenic epilepsy variants. In GEL we found that an increased PRS was associated with higher epilepsy risk, whereas a decreased PRS was protective in people carrying a pathogenic variant. This is in line with a recent family-based study, which found that PRS was associated with increased epilepsy risk and severity among family members carrying the same pathogenic variant.^6^ Similarly, a recent study on hypertrophic cardiomyopathy demonstrated that PRS influenced rare variant penetrance and adverse outcomes, suggesting that the same interplay between rare and common variants might generalize to other diseases. In contrast to GEL, we observed a lower PRS in people with epilepsy and a pathogenic variant in the UKB compared to people without a pathogenic variant, and also compared with those with a variant but without epilepsy. This discrepancy may reflect differences in ascertainment bias between the two cohorts. GEL specifically recruited people with early-onset and syndromic epilepsy such as DEE, which could explain why 13/14 epilepsy cases with a pathogenic variant had severe epilepsies. In contrast, we found no severe epilepsy cases in the UKB, which might be related to inclusion at 40-69 years of age and screening for capacity to understand informed consent, which likely excluded individuals with intellectual disability. Additionally, participation bias in the UKB may have led to an overrepresentation of individuals with above-average intelligence and health outcomes.^24^ Consequently, severe epilepsy cases were likely excluded in UKB and those remaining are most likely milder. The observed low PRS in UKB cases with a pathogenic variant suggest that a pathogenic variant could be sufficient to cause mild forms of epilepsy, while a combination between a pathogenic variant and a high PRS might result in severe epilepsies like DEE.

Our results should be interpreted in the light of several limitations. Due to the rare nature of individual pathogenic variants, we were not able to calculate penetrance estimates per specific variants. To increase robustness and statistical power, we grouped all variants in DEE genes together to assess average penetrance and influence of PRS. However, it is likely that different variants in different genes have a different penetrance. We defined epilepsy variants based on a curated list of DEE genes and established pathogenic epilepsy variants from the ClinVar database, which uses rigorous American College of Medical Genetics (ACMG) criteria for classification. However, we acknowledge that databases like ClinVar are not immune to errors, and some variants may later be reclassified as benign as new evidence emerges.^25^ Variant misclassification could have resulted in decreased penetrance estimates. Additionally, we note that the GEL and UKB have a heterogeneity in phenotyping methods. Cases in the GEL cohort were diagnosed by neurologists using strictly defined criteria. However, epilepsy diagnosis in the UKB is limited to retrospective ICD codes, which is prone to misclassification. The UKB recruitment of individuals aged 40-69 may have limited the ability to capture childhood-onset diseases. For example, childhood-onset epilepsy that goes into remission by adulthood might have been missed at the time of inclusion. Conversely, there could be false-positive labelling of an epilepsy ICD code in cases with provoked seizures and other paroxysmal events. This is in line with previous GWAS and PRS studies using ICD codes to classify epilepsy, which show a strong dilution of signal compared to clinical epilepsy cohorts.^19,21^

In summary, while our study has inherent limitations, it suggests that the penetrance of pathogenic epilepsy variants is lower than previously assumed and influenced by polygenic background. These findings have implications for clinical variant interpretation and underscore the importance of integrating common genetic variation into the interpretation of variant pathogenicity and risk assessment. We hope this work encourages further studies to refine gene-and variant-specific penetrance estimates and unravel how the full spectrum of genetic variation shapes epilepsy risk and severity. Furthermore, our findings may extend to other disorders in which both rare and common variants contribute to disease susceptibility, underscoring the broader relevance of integrating comprehensive genetic risk assessment into clinical practice.

## Acknowledgements

This research was made possible through access to data in the National Genomic Research Library, which is managed by Genomics England Limited (a wholly owned company of the Department of Health and Social Care). The National Genomic Research Library holds data provided by patients and collected by the NHS as part of their care and data collected as part of their participation in research. The National Genomic Research Library is funded by the National Institute for Health Research and NHS England. The Wellcome Trust, Cancer Research UK and the Medical Research Council have also funded research infrastructure. This research has been conducted using data from UK Biobank, a major biomedical database, under application number 199252. We thank the participants of UK Biobank and Genomics England for their invaluable contributions, and the research teams for their work in collecting, processing, and providing access to the data.

## Ethical publication statement

We confirm that we have read the Journal’s position on issues involved in ethical publication and affirm that this report is consistent with those guidelines

