## Supplementary materials for "Penetrance of pathogenic epilepsy variants is low and shaped by common genetic background"

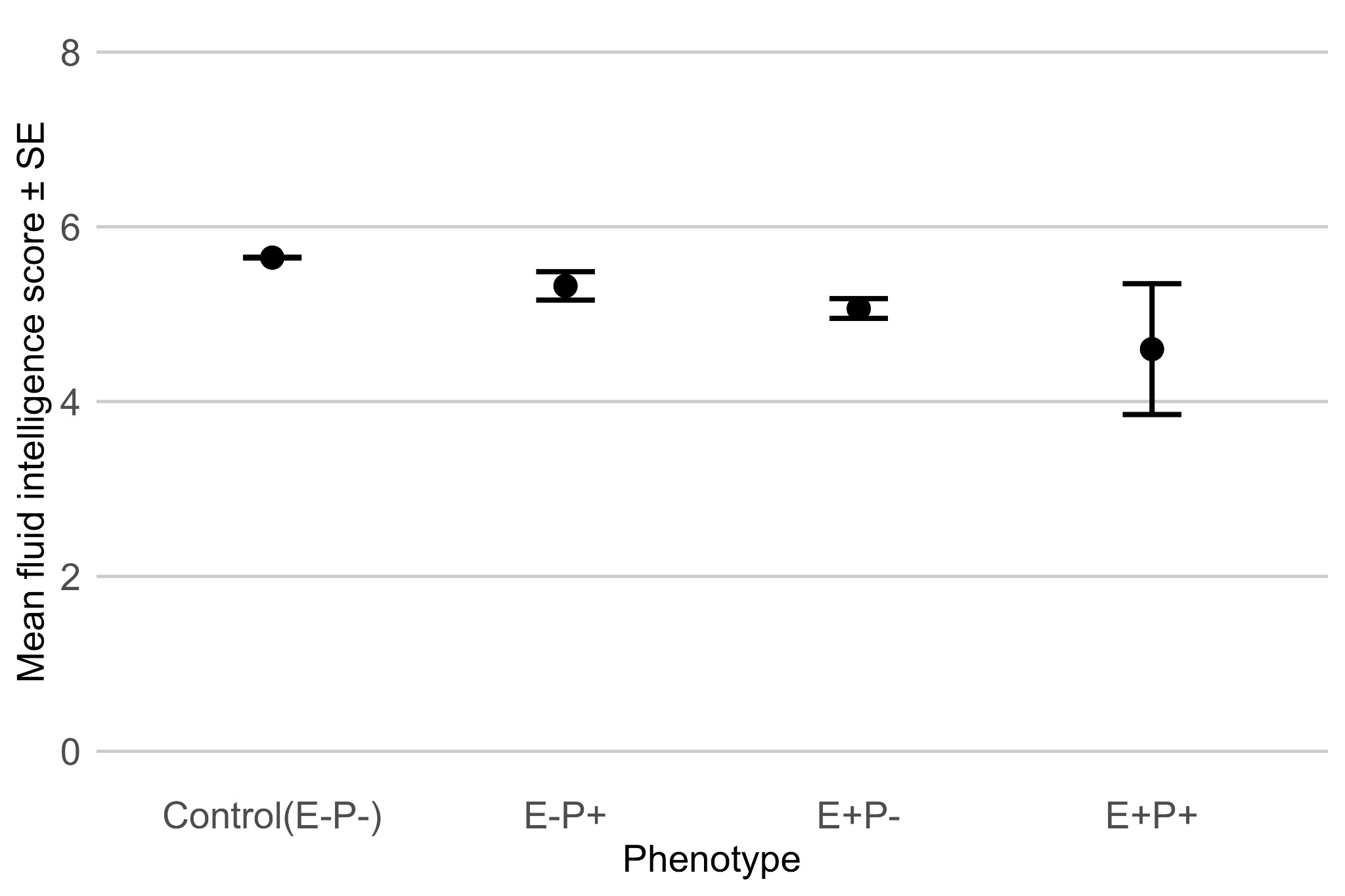

**Supplementary Figure 1: Comparison of fluid intelligence between people with and without epilepsy (E), stratified by pathogenic variant status (P).** Average scores on fluid intelligence tests (range 0-13) conducted in the UKB are displayed per group with standard errors as whiskers. Intelligence scores were available for around half of all subjects (Control: 177,444, E-P+: 136, E+P-: 310, E+P+: 5 subjects).

| Chr | Pos | Ref | Alt | Subjects | Epilepsy/no epilepsy | Gene | RSid | Molecular consequence |
| --- | --- | --- | --- | --- | --- | --- | --- | --- |
| 1 | 196258350 | G | A | 2 | 0/2 | KCNT2 | . | nonsense_variant_variant |
| 2 | 165331569 | G | A | 1 | 1/0 | SCN2A | 1698674881 | splice_donor_variant |
| 2 | 165374749 | T | A | 4 | 0/4 | SCN2A | 1483179196 | missense_variant |
| 2 | 165996069 | T | G | 1 | 0/1 | SCN1A | 1690003122 | missense_variant |
| 2 | 166002491 | T | C | 1 | 0/1 | SCN1A | 121917913 | missense_variant |
| 2 | 166041277 | T | A | 1 | 0/1 | SCN1A | 121918782 | missense_variant |
| 2 | 166045215 | CT | C | 1 | 1/0 | SCN1A | 1697667767 | frameshift_variant |
| 2 | 166046961 | C | T | 1 | 1/0 | SCN1A | . | missense_variant |
| 3 | 11017410 | G | A | 1 | 0/1 | SLC6A1 | 1479789276 | missense_variant |
| 3 | 192335434 | C | T | 1 | 1/0 | FGF12 | 886039903 | missense_variant |
| 9 | 128574726 | C | T | 1 | 0/1 | SPTAN1 | 1851109762 | nonsense_variant |
| 9 | 128626481 | C | T | 2 | 0/2 | SPTAN1 | 1193718145 | missense_variant |
| 9 | 135759686 | G | A | 1 | 1/0 | KCNT1 | 587777264 | missense_variant |
| 12 | 13563850 | G | A | 1 | 0/1 | GRIN2B | 1320154351 | missense_variant |
| 12 | 13615145 | G | T | 1 | 0/1 | GRIN2B | . | missense_variant |
| 12 | 51706586 | ACT | A | 1 | 0/1 | SCN8A | 2138750473 | nonsense_variant |
| 15 | 60531773 | C | G | 1 | 0/1 | RORA | 1555427498 | missense_variant |
| 15 | 92956510 | C | T | 1 | 1/0 | CHD2 | 1064795815 | missense_variant |
| 15 | 92985580 | ACT | A | 1 | 1/0 | CHD2 | 2141851983 | nonsense_variant |
| 15 | 93020132 | G | A | 3 | 0/3 | CHD2 | 150951454 | missense_variant |
| 15 | 93020173 | C | T | 1 | 1/0 | CHD2 | 761127171 | nonsense_variant |
| 16 | 30980736 | CAG | C | 2 | 1/1 | SETD1A | 755127868 | splice_acceptor_variant |
| 19 | 13212719 | G | A | 1 | 0/1 | CACNA1A | . | nonsense_variant |
| 19 | 13228758 | G | A | 1 | 0/1 | CACNA1A | 1296262946 | nonsense_variant |
| 19 | 13228797 | T | C | 1 | 0/1 | CACNA1A | . | splice_acceptor_variant |
| 19 | 13262835 | T | C | 1 | 0/1 | CACNA1A | 1057524483 | splice_acceptor_variant |
| 19 | 13298983 | C | CCCAGGGCCTCCGTGCGT | 1 | 0/1 | CACNA1A | . | frameshift_variant |
| 19 | 13330243 | C | T | 1 | 0/1 | CACNA1A | 2145114772 | splice_donor_variant |
| 19 | 13359652 | ACAGT | A | 1 | 0/1 | CACNA1A | 1599276830 | frameshift_variant |
| 20 | 63413555 | C | T | 1 | 0/1 | KCNQ2 | 118192234 | missense_variant |
| 20 | 63438651 | G | A | 1 | 1/0 | KCNQ2 | 118192215 | missense_variant |
| 20 | 63444762 | G | A | 1 | 1/0 | KCNQ2 | 118192199 | missense_variant |
| 22 | 31766584 | G | A | 1 | 1/0 | DEPDC5 | 1556526609 | splice_acceptor_variant |
| 22 | 31792091 | CA | C | 1 | 1/0 | DEPDC5 | . | frameshift_variant |
| 22 | 31806168 | C | T | 1 | 1/0 | DEPDC5 | 757511744 | nonsense_variant |
| 22 | 31819114 | C | T | 1 | 0/1 | DEPDC5 | 886039263 | nonsense_variant |
| 22 | 31845042 | G | A | 1 | 0/1 | DEPDC5 | 2091643419 | nonsense_variant |

**Supplementary Table 1: List of epilepsy variants found in the GEL cohort.** Chr: chromosome position, Pos: chromosomal position (in GRCh38 assembly), Ref: reference allele, Alt: alternative allele, Subjects: number of GEL subjects carrying the variant, Gene: gene symbol, RSid: reference variant ID from dbSNP (if catalogued), molecular consequence: molecular consequence of the main transcript that is representative of biology at that locus (‘MANE select’) according to ClinVar.

| Chr | Pos | Ref | Alt | Subjects | Epilepsy/no epilepsy | Gene | RSid | Molecular_consequence |
| --- | --- | --- | --- | --- | --- | --- | --- | --- |
| 1 | 1806503 | A | T | 1 | 0/1 | GNB1 | 752746786 | missense_variant |
| 1 | 110673447 | T | C | 2 | 0/2 | KCNA3 | 2100949783 | missense_variant |
| 1 | 110674778 | G | C | 1 | 0/1 | KCNA3 | . | missense_variant |
| 1 | 196258350 | G | A | 10 | 0/10 | KCNT2 | . | nonsense_variant |
| 1 | 244863584 | CCCGCCGCGCGCAACGTAC AACGCAGCACTCACCCGCC GCCGGAGCCCCGGGGCGA | C | 2 | 0/2 | HNRNPU | . | splice_donor_variant |
| 2 | 165091147 | T | C | 1 | 0/1 | SCN3A | 2105621335 | missense_variant |
| 2 | 165308795 | G | A | 2 | 0/2 | SCN2A | . | splice_donor_variant |
| 2 | 165310406 | G | A | 1 | 0/1 | SCN2A | 1057520413 | missense_variant |
| 2 | 165323303 | C | T | 1 | 0/1 | SCN2A | 746060762 | nonsense_variant |
| 2 | 165331328 | A | G | 1 | 0/1 | SCN2A | 2105293230 | splice_acceptor_variant |
| 2 | 165373331 | G | A | 1 | 0/1 | SCN2A | . | missense_variant |
| 2 | 165374724 | A | G | 1 | 0/1 | SCN2A | . | missense_variant |
| 2 | 165374749 | T | A | 35 | 0/35 | SCN2A | 1483179196 | missense_variant |
| 2 | 165991601 | G | A | 1 | 0/1 | SCN1A | 794726739 | nonsense_variant |
| 2 | 165991619 | G | A | 1 | 0/1 | SCN1A | 779614747 | nonsense_variant |
| 2 | 165991799 | C | A | 1 | 0/1 | SCN1A | 1553520107 | nonsense_variant |
| 2 | 165992212 | C | A | 1 | 0/1 | SCN1A | 794726851 | missense_variant |
| 2 | 165992222 | C | T | 1 | 0/1 | SCN1A | . | missense_variant |
| 2 | 165992258 | T | C | 1 | 0/1 | SCN1A | 375651364 | missense_variant |
| 2 | 165992332 | C | T | 2 | 1/1 | SCN1A | 121918622 | missense_variant |
| 2 | 165992360 | G | A | 1 | 0/1 | SCN1A | . | missense_variant |
| 2 | 165994322 | A | G | 2 | 0/2 | SCN1A | 1689707865 | missense_variant |
| 2 | 165999759 | C | T | 1 | 0/1 | SCN1A | 794726699 | nonsense_variant |
| 2 | 166002491 | T | C | 7 | 0/7 | SCN1A | 121917913 | missense_variant |
| 2 | 166039437 | G | A | 1 | 0/1 | SCN1A | 121918784 | missense_variant |
| 2 | 166041277 | T | A | 6 | 0/6 | SCN1A | 121918782 | missense_variant |
| 2 | 166046963 | G | A | 30 | 0/30 | SCN1A | 759121197 | missense_variant |
| 2 | 166048932 | C | A | 1 | 0/1 | SCN1A | . | nonsense_variant |
| 2 | 166051928 | A | G | 1 | 0/1 | SCN1A | 121918780 | missense_variant |
| 2 | 166054638 | G | A | 1 | 0/1 | SCN1A | 1553551312 | missense_variant |
| 2 | 166056467 | C | G | 1 | 0/1 | SCN1A | 1699149959 | missense_variant |
| 3 | 11017342 | G | A | 1 | 0/1 | SLC6A1 | 794726859 | missense_variant |
| 3 | 11017410 | G | A | 1 | 0/1 | SLC6A1 | 1479789276 | missense_variant |
| 5 | 45267103 | C | T | 1 | 0/1 | HCN1 | 1561081319 | missense_variant |
| 5 | 45396551 | C | T | 1 | 0/1 | HCN1 | 1561139569 | missense_variant |
| 8 | 23004537 | G | T | 1 | 0/1 | RHOBTB2 | . | missense_variant |
| 8 | 23006057 | C | T | 1 | 0/1 | RHOBTB2 | 1585190351 | nonsense_variant |
| 9 | 74665521 | G | A | 2 | 0/2 | RORB | 1824250656 | missense_variant |
| 9 | 98454216 | G | A | 1 | 0/1 | GABBR2 | . | missense_variant |
| 9 | 127612443 | A | C | 4 | 0/4 | STXBP1 | 796053379 | intron_variant |
| 9 | 127661206 | G | A | 1 | 0/1 | STXBP1 | . | splice_donor_variant |
| 9 | 127668102 | G | T | 1 | 0/1 | STXBP1 | 1564352002 | nonsense_variant |
| 9 | 127668132 | G | A | 2 | 2/0 | STXBP1 | 587777310 | missense_variant |
| 9 | 128574726 | C | T | 1 | 0/1 | SPTAN1 | 1851109762 | nonsense_variant |
| 9 | 128574777 | C | T | 1 | 0/1 | SPTAN1 | . | nonsense_variant |
| 9 | 128626481 | C | T | 11 | 0/11 | SPTAN1 | 1193718145 | missense_variant |
| 9 | 128632218 | A | G | 2 | 0/2 | SPTAN1 | 1441152520 | missense_variant |
| 9 | 135772718 | C | T | 7 | 0/7 | KCNT1 | 1832838063 | missense_variant |
| 9 | 135777447 | C | T | 1 | 0/1 | KCNT1 | . | missense_variant |
| 11 | 17779489 | C | T | 5 | 0/5 | KCNC1 | 1485166517 | missense_variant |
| 12 | 13615141 | C | T | 1 | 0/1 | GRIN2B | . | missense_variant |
| 12 | 51701142 | A | G | 1 | 0/1 | SCN8A | . | splice_acceptor_variant |
| 12 | 75050795 | C | T | 1 | 0/1 | KCNC2 | . | missense_variant |
| 14 | 62950523 | G | A | 1 | 1/0 | KCNH5 | 1164997707 | missense_variant |
| 15 | 26583406 | G | A | 1 | 0/1 | GABRB3 | 2140737885 | missense_variant |
| 15 | 26621444 | G | A | 1 | 0/1 | GABRB3 | 942355738 | nonsense_variant |
| 15 | 26943239 | C | T | 1 | 0/1 | GABRA5 | 1595438268 | missense_variant |
| 15 | 92956527 | CTCTT | C | 1 | 0/1 | CHD2 | 2141816549 | frameshift_variant |
| 15 | 92972337 | C | T | 1 | 0/1 | CHD2 | 146691368 | nonsense_variant |
| 15 | 93002203 | G | GA | 3 | 0/3 | CHD2 | 749969667 | frameshift_variant |
| 15 | 93020132 | G | A | 37 | 0/37 | CHD2 | 150951454 | missense_variant |
| 16 | 30966890 | C | T | 1 | 0/1 | SETD1A | 2143509708 | nonsense_variant |
| 16 | 30980736 | CAG | C | 1 | 0/1 | SETD1A | 755127868 | splice_acceptor_variant |
| 16 | 30993183 | G | A | 2 | 0/2 | STX1B | 780843272 | nonsense_variant |
| 16 | 30997060 | C | T | 1 | 0/1 | STX1B | . | splice_acceptor_variant |
| 16 | 56336763 | G | A | 1 | 0/1 | GNAO1 | 797044878 | missense_variant |
| 16 | 56351417 | T | TC | 1 | 0/1 | GNAO1 | 2037920369 | frameshift_variant |
| 18 | 57607170 | C | A | 1 | 0/1 | NARS1 | 2122429938 | missense_variant |
| 19 | 13209434 | AGGCGTCG | A | 1 | 0/1 | CACNA1A | 2144506286 | frameshift_variant |
| 19 | 13212218 | T | G | 1 | 0/1 | CACNA1A | 1064796709 | splice_acceptor_variant |
| 19 | 13235648 | C | T | 1 | 0/1 | CACNA1A | 2055849544 | missense_variant |
| 19 | 13235694 | G | A | 1 | 0/1 | CACNA1A | 1555738369 | nonsense_variant |
| 19 | 13235703 | G | A | 3 | 0/3 | CACNA1A | 779576853 | missense_variant |
| 19 | 13262750 | C | T | 2 | 0/2 | CACNA1A | . | missense_variant |
| 19 | 13262772 | G | A | 1 | 0/1 | CACNA1A | 1568473171 | nonsense_variant |
| 19 | 13262781 | G | A | 2 | 0/2 | CACNA1A | 1064794858 | missense_variant |
| 19 | 13262835 | T | C | 4 | 0/4 | CACNA1A | 1057524483 | splice_acceptor_variant |
| 19 | 13286523 | A | AG | 1 | 0/1 | CACNA1A | 757953057 | frameshift_variant |
| 19 | 13303574 | T | G | 4 | 0/4 | CACNA1A | 1555757432 | missense_variant |
| 19 | 13317168 | G | A | 3 | 0/3 | CACNA1A | 121908240 | missense_variant |
| 19 | 13330243 | C | T | 1 | 0/1 | CACNA1A | 2145114772 | splice_donor_variant |
| 19 | 13359635 | T | G | 1 | 0/1 | CACNA1A | . | missense_variant |
| 19 | 13371727 | G | A | 1 | 0/1 | CACNA1A | 886042230 | nonsense_variant |
| 19 | 13371744 | C | T | 1 | 0/1 | CACNA1A | 121908211 | missense_variant |
| 19 | 41986207 | G | A | 1 | 0/1 | ATP1A3 | 2075284959 | missense_variant |
| 20 | 63414101 | T | C | 3 | 0/3 | KCNQ2 | 2145547478 | missense_variant |
| 20 | 63438651 | G | A | 1 | 1/0 | KCNQ2 | 118192215 | missense_variant |
| 20 | 63438690 | C | T | 1 | 0/1 | KCNQ2 | 2145712541 | missense_variant |
| 20 | 63439608 | G | A | 1 | 0/1 | KCNQ2 | 864321707 | missense_variant |
| 20 | 63439651 | G | A | 1 | 0/1 | KCNQ2 | . | missense_variant |
| 20 | 63439718 | C | T | 9 | 0/9 | KCNQ2 | 375363057 | intron_variant |
| 20 | 63442518 | G | C | 1 | 0/1 | KCNQ2 | . | missense_variant |
| 20 | 63444729 | C | T | 1 | 1/0 | KCNQ2 | 118192200 | missense_variant |
| 20 | 63444834 | T | C | 1 | 0/1 | KCNQ2 | 1555873823 | missense_variant |
| 20 | 63472462 | A | G | 1 | 0/1 | KCNQ2 | 118192186 | missense_initiator_codon_variant |
| 20 | 63490687 | G | A | 1 | 0/1 | EEF1A2 | . | missense_variant |
| 21 | 43417680 | G | T | 3 | 0/3 | SIK1 | . | nonsense_variant |
| 22 | 31754942 | C | G | 1 | 0/1 | DEPDC5 | 768241563 | nonsense_variant |
| 22 | 31783940 | ATATT | A | 1 | 0/1 | DEPDC5 | . | frameshift_variant |
| 22 | 31809648 | G | T | 1 | 0/1 | DEPDC5 | 2088011240 | splice_donor_variant |
| 22 | 31810562 | C | T | 1 | 1/0 | DEPDC5 | 2148707379 | nonsense_variant |
| 22 | 31815005 | C | T | 5 | 0/5 | DEPDC5 | 587777459 | nonsense_variant |
| 22 | 31815005 | CGA | C | 2 | 0/2 | DEPDC5 | . | frameshift_variant |
| 22 | 31819054 | C | T | 1 | 0/1 | DEPDC5 | . | nonsense_variant |
| 22 | 31819114 | C | T | 1 | 0/1 | DEPDC5 | 886039263 | nonsense_variant |
| 22 | 31845042 | G | A | 1 | 1/0 | DEPDC5 | 2091643419 | nonsense_variant |
| 22 | 31870695 | C | T | 1 | 1/0 | DEPDC5 | 2092816377 | nonsense_variant |
| 22 | 31873333 | G | A | 2 | 0/2 | DEPDC5 | 1261611694 | splice_donor_variant |
| 22 | 31874286 | CT | C | 1 | 0/1 | DEPDC5 | 1309891064 | frameshift_variant |
| 22 | 31876262 | C | T | 1 | 0/1 | DEPDC5 | 886039268 | nonsense_variant |
| 22 | 31879713 | C | T | 1 | 0/1 | DEPDC5 | 886039269 | nonsense_variant |

**Supplementary Table 2: List of epilepsy variants found in the UKB cohort.** Chr: chromosome position, Pos: chromosomal position (in GRCh38 assembly), Ref: reference allele, Alt: alternative allele, Subjects: number of UKB subjects carrying the variant, Gene: gene symbol, RSid: reference variant ID from dbSNP (if catalogued), molecular consequence: molecular consequence of the main transcript that is representative of biology at that locus (‘MANE select’) according to ClinVar.

| Group comparison | Odds ratio | P-value |
| --- | --- | --- |
| E+P+ vs E-P+ | 1.84 | 0.047 |
| E-P- vs E-P+ | 0.72 | 0.069 |
| E+P+ vs E+P- | 0.80 | 0.388 |
| E+P+ vs E-P- | 1.91 | 0.019 |
| E+P- vs E-P- | 1.43 | <2e-16 |

**Supplementary Table 3: Statistical tests of group comparisons in GEL.** Epilepsy PRS was compared between people with and without epilepsy (E), stratified by pathogenic variant status (P). Odds ratios and P-values of logistic regressions to compare each group are displayed.

| Group comparison | Odds ratio | P-value |
| --- | --- | --- |
| E+P+ vs E-P+ | 0.35 | 0.086 |
| E-P- vs E-P+ | 0.87 | 0.121 |
| E+P+ vs E+P- | 0.20 | 0.007 |
| E+P+ vs E-P- | 0.33 | 0.032 |
| E+P- vs E-P- | 1.46 | 1.67e-11 |

**Supplementary Table 4: Statistical tests of group comparisons in UKB.** Epilepsy PRS was compared between people with and without epilepsy (E), stratified by pathogenic variant status (P). Odds ratios and P-values of logistic regressions to compare each group are displayed.
